# Rapid threat detection in SARS-CoV-2

**DOI:** 10.1101/2022.08.05.22278480

**Authors:** Christopher L. Barrett, Fenix W.D. Huang, Thomas J.X. Li, Andrew S. Warren, Christian M. Reidys

**Affiliations:** Biocomplexity Institute and Initiative, University of Virginia, Charlottesville, VA, USA; Department of Computer Science, University of Virginia, Charlottesville, VA, USA; Department of Mathematics, University of Virginia, Charlottesville, VA, USA

## Abstract

This paper presents a novel virus surveillance framework, completely independent of phylogeny-based methods. The framework issues timely alerts with an accuracy exceeding 85% that are based on the co-evolutionary relations between sites of the viral multiple sequence array (MSA). This set of relations is formalized via a motif complex, whose dynamics contains key information about the emergence of viral threats without the referencing of strain prevalence. Our notion of threat is centered at the emergence of a certain type of critical cluster consisting of key co-evolving sites. We present three case studies, based on GISAID data from UK, US and New York, where we perform our surveillance. We alert on May 16, 2022, based on GISAID data from New York, to a critical cluster of co-evolving sites mapping to the Pango-designation, BA.5. The alert specifies a cluster of seven genomic sites, one of which exhibits D3N on the M (membrane) protein–the distinguishing mutation of BA.5, three encoding ORF6:D61L and the remaining three exhibiting the synonymous mutations C26858T, C27889T and A27259C. New insight is obtained: when projected onto sequences, this cluster splits into two, mutually exclusive blocks of co-evolving sites (m:D3N,nuc:C27889T) linked to the five reverse mutations (nuc:C26858T,nuc:A27259C,ORF6:D61L). We furthermore provide an in depth analysis of all major signaled threats, during which we discover a specific signature concerning linked reverse mutation in the critical cluster.

## 1 Introduction

### Background

During the COVID-19 pandemic, caused by the SARS-CoV-2 viruses [1], many variants were recognized to pose a threat, in particular Alpha, Delta, Omicron including its sub-lineages BA.2, BA.2.12, BA2.12.1, BA.4 and BA.5 [2–5]. The virus has infected hundreds of millions of people and taken the lives of million others worldwide. Furthermore, this virus exhibits a significant increase in transmissibility, due to the constantly evolving spike protein region. This mediates the binding efficacy between the viral spike protein and the human ACE2 receptor [6, 7] and thus the efficiency with which the virus enters a new cell. Viral mutations not only impact the transmission rate, but also lead to increasing reinfection cases [8], raising questions regarding the efficiency of vaccines. Closely monitor the evolution of this virus is critical to better understand and predict the course of the pandemic.

Current SARS-CoV-2 genomic surveillance [9–15] employs the GISAID database [10], which promotes the immediate sharing of more than 10 million SARS-CoV-2 genomic sequences with temporal and geographic information. Resources such as Nextstrain, *CoVariants* and Pango [11, 13–15] utilize phylogenetic analysis of viral genomes to track and classify sequenced samples throughout the world [16–20]. To this end, sequences are organized in multiple sequence alignments (MSAs) [21] and a tree structure is inferred from the similarities between sequence pairs [22]. A leaf node in this derived tree represents a genomic sequence and the length of the path connecting two leaf nodes represents the estimated time of divergence from their common ancestor. In the Pango system, a monophyletic cluster of sequences is manually designated as a lineage if the clade they represent is sufficiently close and well represented [15]. Pango [15] is based on worldwide data and generated on April 5, 2022 (Issue#517) based on seventy of nine sequences from *South Africa*. This led to the designation of the lineages BA.4 and BA.5 on April 7. In order to put the frequency of Issues and lineage designation into context, on average Pango generates one Issue as well as one lineage per day.

In phylogeny-based methods, mutations, which are commonly found in a lineage (over 75% [12]) are considered to be its characteristics. Characteristic mutations are the input of subsequent biological analysis, however, these are not always disjoint across lineages. For instance, the D614G mutation on the spike protein of the SARS-CoV-2 genome is commonly present in most of the variants [23–25], even though some of them are distant in the phylogenetic tree. While the D614G mutation is relevant in the context of evaluating adaption relative to the wild type strain, it arguably becomes less and less relevant when comparing adaptation of variants distant from the wild type. Tools like *Covariants* provide a general picture on how characteristic mutations are distributed over different variants. The appearance and disappearance of these characteristic mutations indicates that there are intrinsic relationships among these mutations that are difficult to express within this framework.

### Novelty

Our surveillance framework differs substantially from phylogeny-based methods. The common ground is that it employs the fact that the observed sequences are the evolutionary “winner”, in order to deduce key information about the system. The categorical difference between ours and any phylogeny-based method, is that we consider genomic sites instead of sequences as the basic building blocks. This can be paraphrased as the “dual” of phylogenetic analysis: instead of considering the relations among rows in an MSA (say, via Hamming distances), we consider the relations among columns, where each column represents a genetic position (a site or locus). This amounts to a holistic interpretation of an MSA. It considers an MSA to be an irreducible object, exhibiting a relational “structure”, analogous to a “molecular consensus structure”, exhibited by an MSA. This MSA-structure is called its *motif complex* and represents the set of co-evolutionary relations among the sites. The idea is that if a group of sites is linked, they are likely to mutate simultaneously in a way that their linkage is maintained.

Co-evolutionary relations among sites are conceptually different from nucleotide patterns. It is these co-evolutionary relations, the “patterns that survive”, that evolve as the virus adapts and viral strains compete within the population. This is analogous to the evolution of bio-physical phenotypes acted upon by selection. This illustrates a key difference between the motif-based versus phylogeny-based variants: selection acts directly on the relational structure that forms the basis for a motif-based variant but only indirectly on mutational patterns. Accordingly, motif-based analysis is well suited for understanding and predicting viral evolution.

With regards to the actual evolution of these co-evolutionary relations, the kinetic folding [26–28] of bio-molecular structures represents an instrumental analogy. In kinetic folding, certain base pairs (binary relations between sites) have to be opened before a new, energetically more optimal configuration can be realized. This requires some “tunneling” as in the interim sub-optimal configurations are assumed. In Section 3 we will demonstrate that the viral population undergoes relational “phase transitions”, representing an analogue of such tunneling. Remarkably, almost every time after these phase transitions, a collection of linked reverse mutations is observed. This indicates that certain relations have to be destroyed, which is facilitated by reversing to the wild type, in order to assume a more optimal relational structure down the line.

### Omicron^P^ is not Omicron^M^

We stipulate that motif-based alerts occur simultaneously with phylogeny/prevalence-based alerts. However, there is a fundamental difference between motif- and phylogeny-based alerts, that is of particular relevance to subsequent biological analysis as well as vaccine design. To illustrate this point, let us consider the transition from Delta to Omicron including BA.1 in the US. The phylogeny-based variant (P-variant), Omicron^P^, is tantamount to a set of 33 defining mutations and the Omicron^P^ sub-lineage BA.1^P^ consists of a set of 26 defining mutations [15]. The BA.1^P^ mutations are assigned to be disjoint to the Omicron^P^ mutations since BA.1^P^ is a direct descendant of Omicron^P^. As such, any sequence containing the Omicron^P^ mutations is considered to be Omicron^P^ and any sequence containing both Omicron^P^ and the BA.1^P^ mutations is considered to be BA.1^P^.

Since any type of meaningful threat detection has to consider the emergence of variants at minuscule prevalence levels, connecting the notions of site and mutation, inevitably recruits some notion of differential. Specifically, a site *e*xhibits a mutation if the proportion of the mutation in the given MSA at that site is monotonously increasing. Note that a site can, in general, simultaneously exhibit multiple mutations as well as a mutation and its reverse mutation. In our case studies, we observe that sites contained in critical clusters almost always exhibit either exclusively mutations or reverse mutations.

We observe that the motif based variant (M-variant), Omicron^M^, represents a unique cluster of 55 co-evolving sites. These sites exhibit 30 Omicron^P^ mutations, 13 BA.1^P^ mutations, and 13 Delta^P^ reverse mutations. The cluster enlarges in the following week to contain 68 sites, which then encompasses all 26 sites exhibiting BA.1^P^ mutations. We claim that these 13 Delta^P^ linked, reverse mutations are as relevant to biological analysis as any of the other mutations. In Fig. 1 we display the linkage between all Omicron^M^ together with BA.1^M^ mutations and Omicron^P^ together with BA.1^P^ mutations on the spike protein in December week 1, 2021, when our framework triggers an alert. We also display the linkages of sites in Delta^M^, BA.2^M^, BA.4^M^ and BA.5^M^ when their corresponding motif-based alerts were triggered. We observe linkages between mutations and reverse mutations in all cases, showing that they are pieces of the puzzle. By abuse of notation, we do not distinguish P- and M-variants when it is clear from the context which one is referred to.

**Fig 1.**
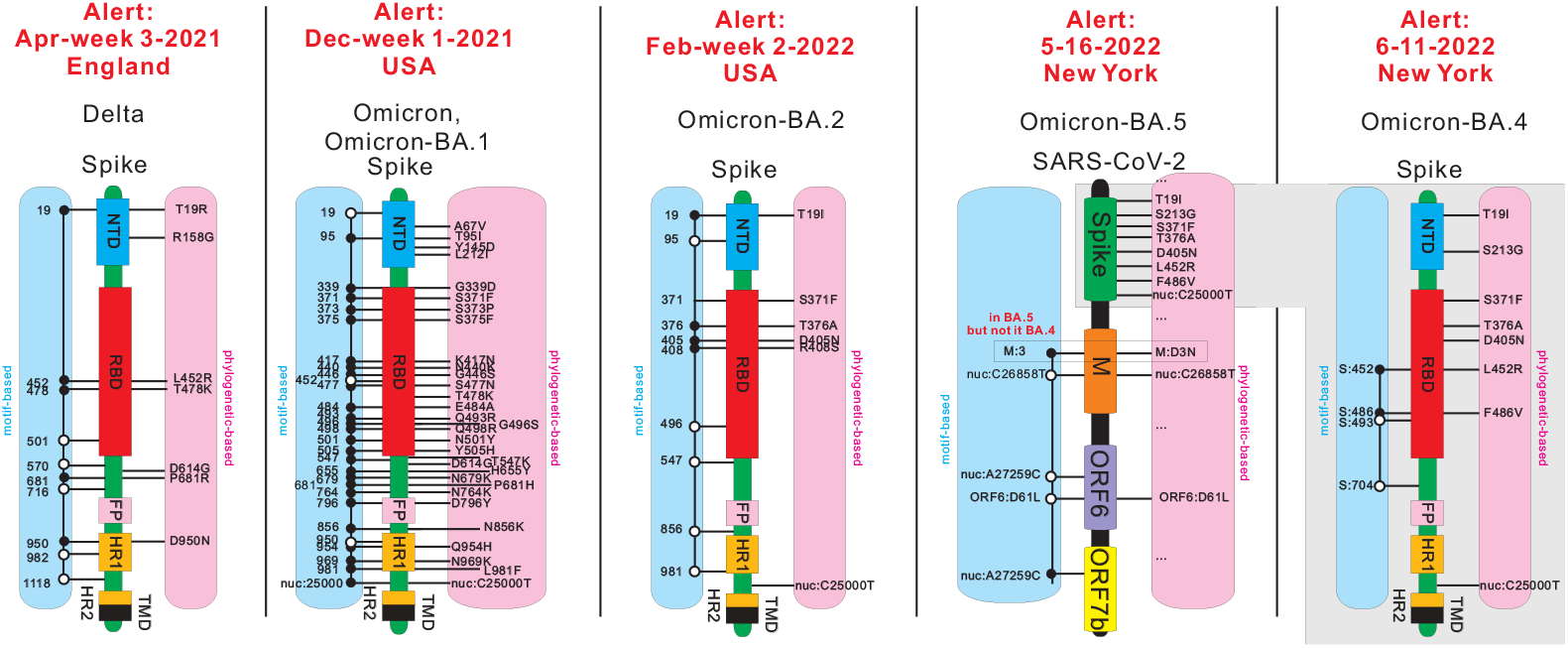
Comparison between linked co-evolving sites in motif-based variants and mutations in phylogeny-based variants. We consider Delta, Omicron together with BA.1, BA.2, BA.4, and BA.5. Except for BA.5, we schematically represent the SARS-CoV-2 spike protein region via functional blocks (NTD:N-terminal domain, RBD:receptor binding domain, FP:fusion peptide, HR 1:heptad repeat 1, HR 2:heptad repeat 2, TMD:transmembrane domain). We then annotate the linked sites of a motif-based variant (M-variant) on the left (blue), and the defining mutations of a phylogeny-based variant (P-variant) on the right (pink). For BA.5, we annotate the sites and mutations using SARS-CoV-2 spike protein, M protein, ORF6 and ORF7b. For any M-variant, sites exhibiting mutations are labeled by black circles and sites exhibiting reverse mutations are labeled by white circles.

Fig. 1 summarizes key observations of this paper concerning the alerts issued in the context of various case studies, detailed in Section 2. It provides a comparative analysis of the phylogeny- and motif-based projections of the virus for virtually all observed alerts. BA.2.12.* is a notable exception, which triggers an alert but does not exhibit linked, reverse mutations. Subject to this exception, all alerts of Fig. 1 exhibit linked, reverse mutations in the underlying critical cluster. Furthermore, BA.5 does not differ from BA.4 w.r.t. spike protein mutations, however, it exhibits a critical cluster, described above, that is independent of any spike protein mutations and involves a distinguished mutation on the M protein.

A key result of this paper is the notion of the differential quantifying the changes in relations within the MSA. The *differential* of a motif complex at time *t*, Δ = Δ(*t*), reflects the degree of change of the motif complex. The differential is critical for our definition of an alert. We are able to devise a surveillance system that alerts to keystone events, including the emergence and disappearance of variants of concern as well as neutral and deleterious variants. The system is tested in the context of viral populations at different spatial and temporal scales: country, state, city and monthly, weekly, 3-day periods. The alerts produced are specific to, but consistent across various geographic locations and are robust with respect to different parameter choices.

While sequences (MSA-rows) can be compared directly using similarity measures, like Hamming distance, the comparison of MSA-columns is not that straightforward. The relations among columns in an MSA were studied using a framework introduced in [29] for the SARS-CoV-2 genome. A simplicial complex, the *motif complex*, is used to model *k*-ary relations among *k* sites. The simplicial complex formalizes a topological enhancement of graph models in which edges represent pairwise relations (*k* = 2). The nucleotide distribution of a column is considered as a random variable with the nucleotide types as its states. The motif complex is represented as a collection of simplices, in which each *k*-simplex captures the *k*-ary correlation among *k* polymorphic sites. These correlations can be understood by linkage disequilibrium [30] and the evolutionary distance introduced in [29, 31].

Studying the differential of motif complexes allows us to understand how the relational structure of the MSA evolves. Via the differentials of motif complexes, we derive a genomic surveillance system whose concept of alert is based on two criteria: (a) a sufficiently large Δ, (b) cluster criticality i.e. persistence of the cluster, in combination with an increasing average entropy over all involved sites.

In case of an alert, the underlying Δ quantifies either the emergence of constellations of mutations, a de novo cluster, or the reorganization of linkages among existing ones. The former manifests in phylogenetic analysis as the emergence of a new lineage and the latter is currently, to our knowledge, not expressed in the context of a lineage/sub-lineage classification scheme. Even though sub-lineages have a similar set of mutations, their underlying mutational relations can be quite different. Lineages that exhibit an increasing average entropy are indicative of the fact that the emerging constellation of mutations is circulating and hence play an essential role in the viral population.

We consider a variant to be a *key variant*, if at a certain location it constitutes at least 50% of the population. At present, Pango has designated over 2000 lineages and sub-lineages [15]. We will show that we issue alerts not much later than lineages are designated. However, alerts identify the emergence and disappearance of key variants with an accuracy of 88.6%, 100% and 99.3% in UK, USA and New York, respectively.

The workflow of our framework is depicted in Fig. 2. We begin with the MSAs of viral sequences at discrete times, see Fig. 2 (A). Following [29], we compute the correlation relations among sites for each MSA in (B), and construct their motif complexes in (C). To measure the differential Δ of motif complexes, we construct the correspondence between their clusters in (D), see Section 4 for details. In (E), we integrate the correspondence to derive Δ and issue alerts.

**Fig 2.**
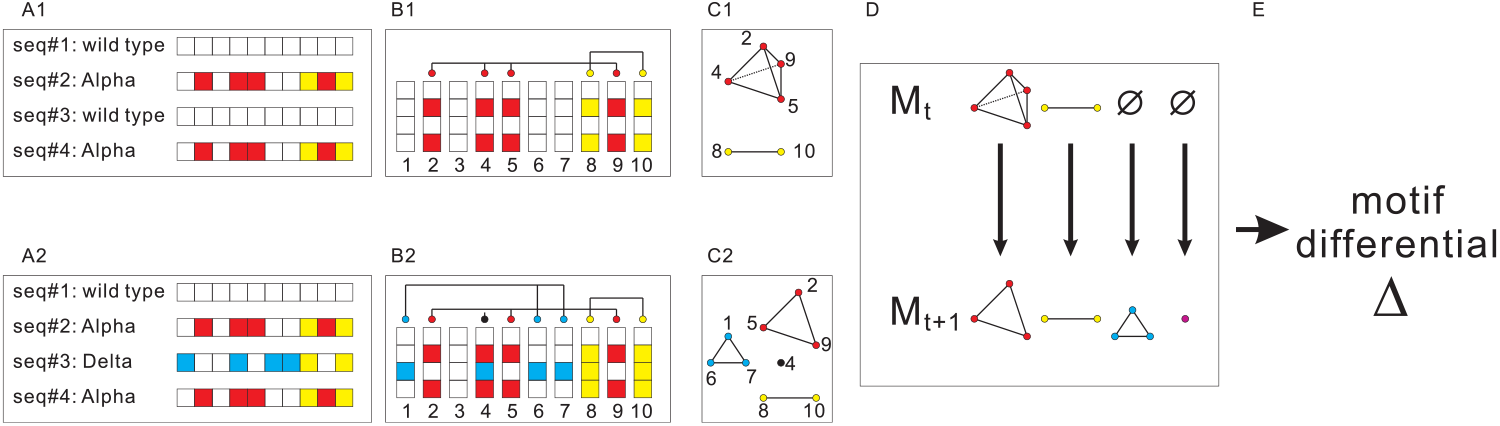
From multiple sequence alignment to alert. A1: A viral population consisting of 2 sequences of the Alpha variant and 2 sequences of the wild type. Boxes (red/yellow) indicate Alpha-mutations relative to the reference sequence. A2: a new variant, Delta, emerges in the population of A1. Boxes (blue/yellow) denote the characteristic mutations of Delta. Here there exists a shared site that exhibits the variant-specific mutations P681H in Alpha and P681R in Delta in the spike protein region. B1 and B2: Correlation relation among the columns of A1 and A2. Columns (sites) in relational patterns are linked. C1: the motif complex of B1: a 4-simplex (red) and a 2-simplex (yellow). C2: the motif complex of B2: two 3-simplices (red/blue), a 2-simplex (yellow), and a 1-simplex (purple). D: computing the derivative: comparing two motif complexes. E: the alert.

The paper is organized as follows: in Section 2 we study the differential. In Sub-Section 2.1, we analyze the differentials of SARS-CoV-2 genomic data in UK, USA, and New York respectively. We perform this analysis during time frames in which key milestone events occur. In Sub-Section 2.2, we provide a consistency analysis of the motif complex and a robustness analysis of key parameters for computing the differential. In Section 3 we integrate and discuss our results. In Section 4 we present the details of our analysis method and protocol. In the Supplementary Materials, we present and analyze specific scenarios that are designed to calibrate the differential.

## 2 Results

Significant changes in the relational structure of the MSA are not random events. Above a specific threshold, these will trigger an alert. Our surveillance system takes an MSA at a specific time as input and computes an approximation of its motif complex, the details of which are given in Section 4. The motif complex in turn gives rise to a collection of clusters of sites that are closely correlated and these allow to quantify how the complex changes. We issue an alert based on two criteria: a) differential (*D*), and b) cluster criticality (*C*).

In Section 4 we detail how to derive differentials Δ = Δ(*t*). Δ quantifies the probability that the emergence of new clusters or the reorganization of existing clusters represents a significant change. We set *D* = 1 if Δ ≥ 5 and *D* = 0, otherwise.

Cluster criticality is based on persistence and average entropy of all involved sites. Persistence reflects the fact that the cluster is present in the population over a sufficiently long period of time, and entropy indicates the diversity of linked sites. Note, the notion of persistence is based on the transition from the cluster configuration at time *t* to the cluster configuration at time *t* + 1, as detailed in Section 4.6. A cluster is considered persistent if it is present for more than five times the sampling time.

Increase of the average entropy reflects the fact that the mutations exhibited by the site are dominating the population. In case average entropy decreases, the linked sites become less diverse i.e. either the exhibited mutations are fading or the domination of the strain is nearly complete. Persistent clusters, whose entropy increases exceeded the threshold parameter 0.35 represent potential threats and we refer to these as *critical*. We set *C* = 1 if at least one critical cluster experiences a significant change in size (*>* 30%) and set *C* = 0, otherwise. An alert is triggered when both of the two criteria are met: (*D* = 1, *C* = 1).

We apply our surveillance system in two ways: retrospectively and prospectively. For the retrospective analysis we can utilize the fact that threats are well identified and can thus be referenced. Moreover variant prevalence is known, which allows to evaluate our surveillance results. However, when performing the surveillance we have no *a priori* knowledge aside from being able to map any site-cluster we discover. The retrospective analysis is performed on sequence data from the UK [10] over a time period covering the emergence of Alpha and Delta and data from the US [10] in the context of the emergence of Omicron and BA.2.

The concurrent/prospective analysis is based on New York data [10] covering the emergence of BA.2.12/BA.2.12.1 and BA.4/BA.5. In this scenario, not all threats can be mapped and are then labeled as *unknown*. The results demonstrate that our surveillance system can accurately alert to the emergence and disappearance of key variants in real time. More importantly, the critical clusters triggering our alerts can succinctly identify relevant mutations of variants. Our results show that variants are not only characterized by the linkage among mutations that deviate from the wild type, but also by the linkage among reverse mutations. The later are usually not factored in, since the current definition of a variant is in reference to the wild type.

In-depth analysis of the motif complex is provided at the end of this section, where we analyze SARS-CoV-2 data from 39 geographic locations at the time of the emergence of Omicron.

### 2.1 Case studies

#### 2.1.1 UK

In order to put the subsequent analysis into context, let us first recall variant prevalence, using Pango [15] lineage designation, see Fig. 3 (A). We observe three key events: firstly, the Alpha variant emerges during November, week 1, 2020, at which point it represents 5% of the population. Secondly, Delta as well as its AY.3 sub-lineages emerge during April, week 2, 2021 (at 5%), and thirdly, Alpha disappears in June week 4, 2021, and the AY.3 sub-lineage dominates (≥ 95%) in June, week 3, 2021.

**Fig 3.**
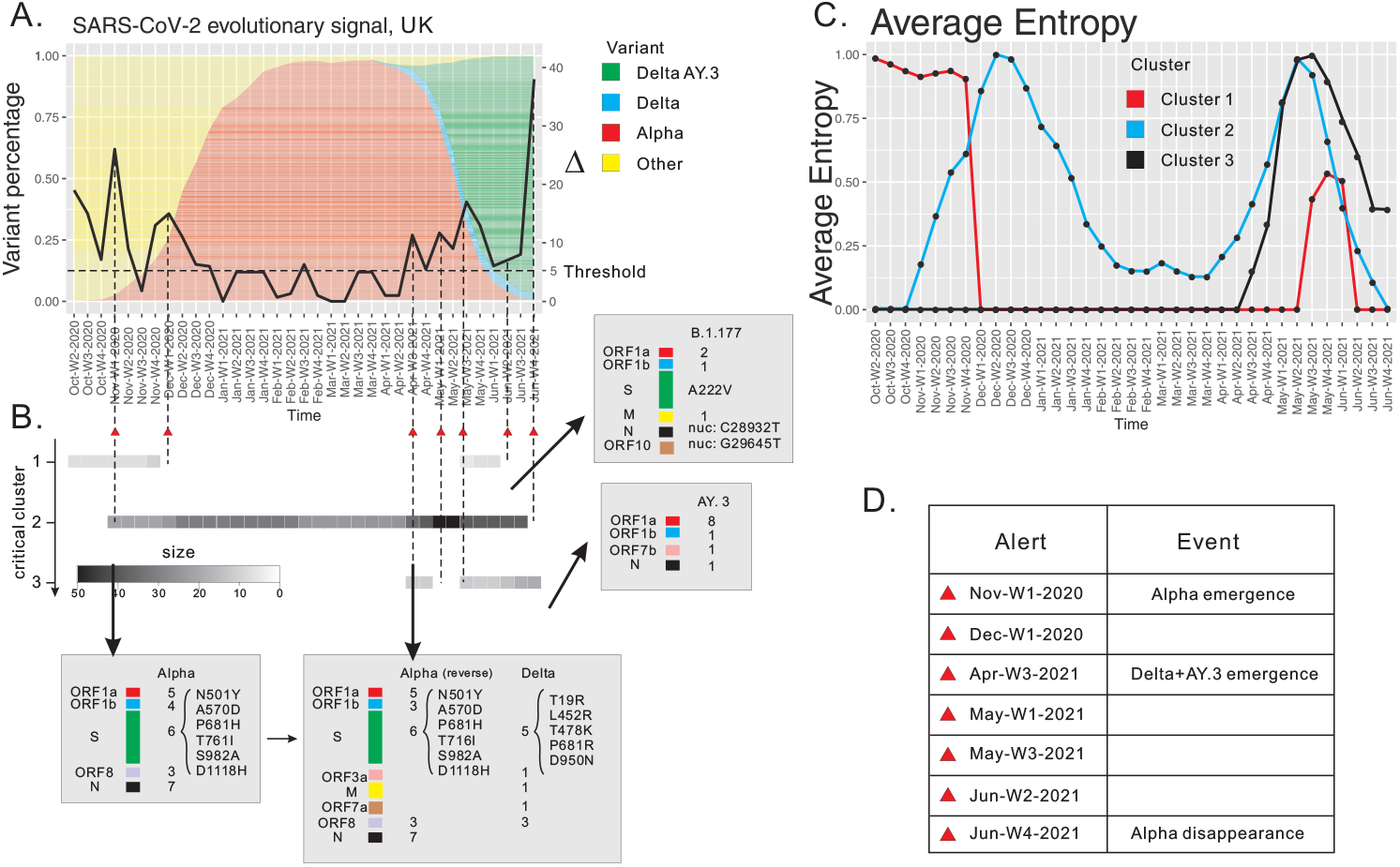
SARS-CoV-2 surveillance based at weekly resolution of UK data, October week 2, 2020 through June week 4, 2021. (A) The differential, displayed as a curve with values on the y-axis. (B) The persistence of critical clusters, represented by bar codes, whose grey scale reflects cluster size. Annotation of sites contained in critical clusters. We cross-reference the sites to characteristic mutations of reported variants. Reverse mutations are distinguished from mutations. Spike protein mutations as well as key mutations reported in the literature are listed. (C) Average entropy of sites contained in the critical cluster. (D) List of alerts (red triangles).

Our framework issues seven alerts, three of which detect the above mentioned three keystone events, see Fig. 3 (A). The three alerts are triggered in November week 1, 2020, April week 3, 2021 and June week 4, 2021, where we identify the emergence of Alpha, of Delta and AY.3, and the disappearance of Alpha, respectively. The remaining four alerts do not directly match the keystone events regarding prevalence, however, they reflect substantial changes in the motif complex. These changes are induced by the reorganization of clusters, that do not manifest in terms of prevalence. In addition, we show that our analysis provides novel insights via M-variants, which enhance the classification based on P-variants. To this end we represent critical clusters as persistent bar codes (grey) in Fig. 3 B.

We identify three critical clusters C1-C3. C1 contains seven sites, one of which exhibits a non-synonymous mutation C22227T, causing the amino acid change A222V in the spike protein [32, 33]. Furthermore, two sites of this cluster exhibit the nucleotide mutations, C28932T and G29645T. All these three mutations are reported as signature mutations of the B.1.177 lineage according to [32]. In Fig. 3 (D) we display the average diversity of the cluster, observing a sharp decrease of the average entropy to zero during December week 1 2020. This indicates that the virus reorganizes itself and changes its co-evolutionary signature. C1 re-emerges during May, week 3 2021, but disappears quickly within the following three weeks. We observe that C1 is related to B.1.177 and purified by subsequent variants.

C2 emerges during December week 1 2020, which triggers our first alert due to the significant change of motif complex. Cross-referencing with Pango lineages, we find that all 25 sites of C2 exhibit mutations among the 28 characteristic mutations of the Alpha variant [14], see Fig. 3 C. One site exhibits the nucleotide mutation A23063T, which causes the amino acid mutation N501Y in the spike protein and is reported as the bio-marker of Alpha [34, 35]. Therefore we conclude that C2 corresponds to the emergence of Alpha when connected to Pango designations. We can infer from our analysis that the Alpha variant is spreading in the viral population, since the average entropy of the cluster is increasing from November week 1, 2020 to December week 2, 2020.

C2 evolves: its size grows to 28 during December week 1 2020, and eventually to 31 in the following week. The sites that are incorporated are C1-sites, i.e. we observe a merger of C1 and C2. The reason of this relation is due to the B.1.177 sites exhibiting linked reverse mutations to the Alpha sites. As Alpha starts to outcompete the previous reigning variant B.1.177, we observe a decrease in the proportion of B.1.177 mutations in the viral population. As a result, the nucleotides in the corresponding sites are reversed to the wild type along the circulation of Alpha. C2 increases again during April, week 3 2021. The additional sites exhibit the defining mutations of the Delta variant (Pango lineage B.1.617.2) and none of these is an AY.3-exclusive site. Furthermore, the Alpha sites exhibit reverse mutations. During the same time period, the average entropy increases, indicating that Delta takes over the population while reversing those mutations whose sites are exclusive to Alpha.

Our observations regarding C2 suggest that the Delta variant is characterized by a cluster of sites, which can be split into two blocks. One block exhibits the constellation of Delta mutations reported by Pango, the other block exhibits linked, reverse mutations, which effectively restore the characteristic mutations of Alpha back to the wild type. It is worth pointing out that the linkage among mutations and reverse mutations is rarely investigated in the current literature. We hypothesize, that the presence of reverse mutations is indicative of the threat potential of a variant and we are currently developing this into an enhanced alert criterion.

C2 disappears in June week 4 2021, triggering an alert, which is due to the disappearance of Alpha and the domination of Delta.

The third critical cluster, C3, emerges during April, week 3, 2021, when Delta sites are merging with C2. C3 contains 11 sites, all of which exhibit AY.3 mutations. Its average entropy is increasing, as AY.3 has increasing prevalence in the population together with Delta.

The motif analysis provides deeper insight into the evolutionary dynamics of the virus as a whole, that, to the best of our knowledge, is not observed via phylogenetic analysis. In particular, the mutation s:P681H in Alpha exhibit a different genetic linkage to s:P681R in Delta, even though they share the same site [36–39]. The reorganization of sites in the motif complex reveals a more detailed picture of the inter-dependencies within the constellation of mutations.

#### 2.1.2 US

Our second retrospective analysis is based on US, GISAID data starting from November week 2, 2021 until March week 1, 2022. This time frame covers the transition from Delta to Omicron. In analogy to our approach in the previous subsection, we display the prevalence of key variants in the US using Pango lineage designation in Fig. 4 (A). We observe three key events: firstly, the Delta variant dominates (95%) from November week 2, 2021 to December week 1, 2021, secondly, Omicron as well as its BA.1 sub-lineage emerge in December week 3, 2021 (5% of the population) and dominate (95%) in January week 3, 2022, and thirdly the Omicron sub-lineage BA.2 emerges in February week 4, 2022 (5%).

**Fig 4.**
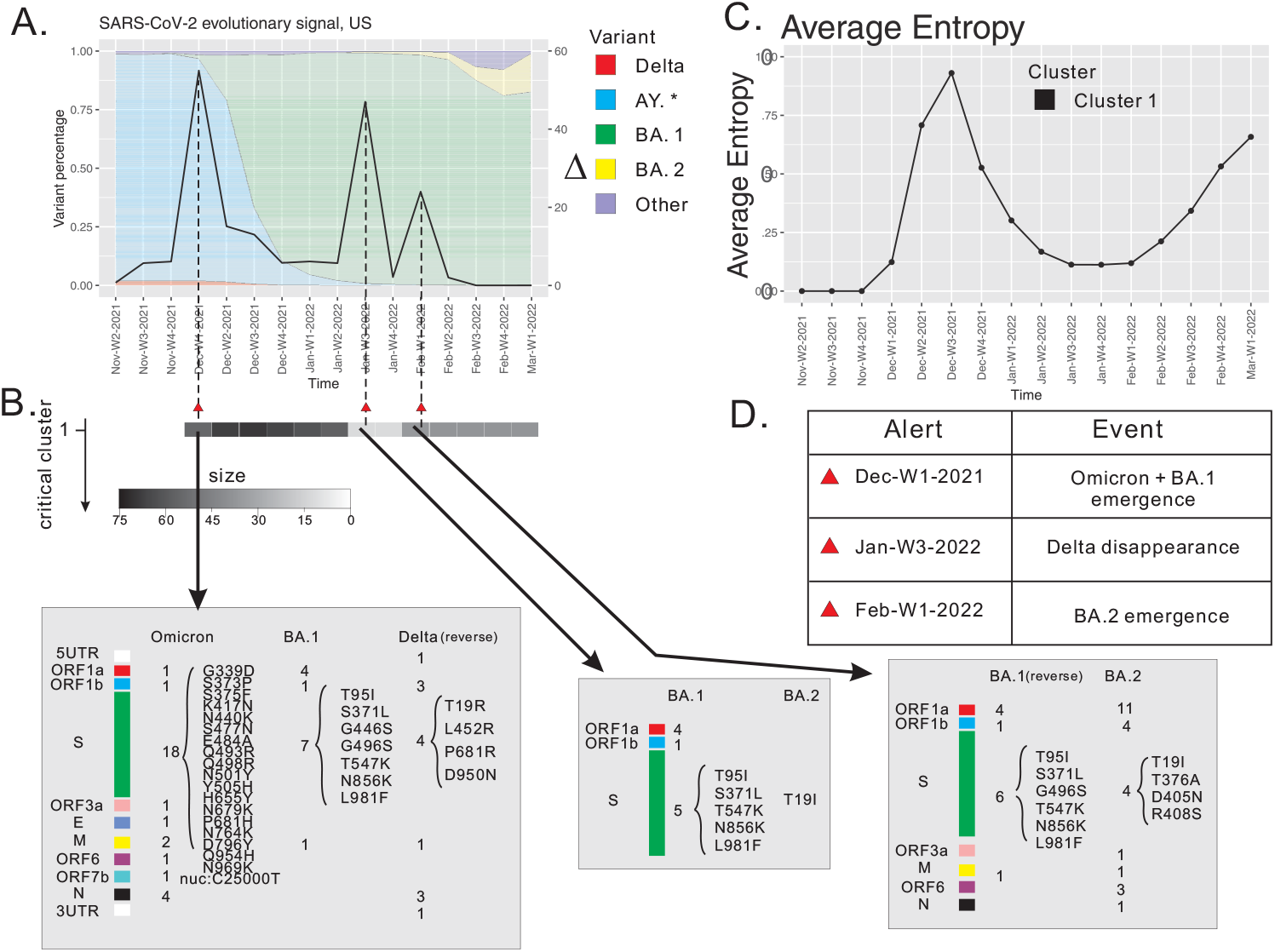
SARS-CoV-2 surveillance based at weekly resolution of USA data, November week 2, 2021 through March week 4, 2022. (A) The differential is displayed as a curve with its values on the y-axis. (B) The persistence of critical clusters, represented by bar codes, whose grey scale reflects the cluster size. Annotation of sites contained in critical clusters. We cross-reference the sites to characteristic mutations of reported variants. Reverse mutations are distinguished from mutations. Spike protein mutations as well as key mutations reported in the literature are listed. (C) Average entropy of sites contained in a critical cluster. (D) List of alerts (red triangles).

Our alert system triggers three alerts: November week 3, 2021, January week 3, 2022, and February week 4, 2022, see Fig. 4 (A). All of them are associated to a unique critical cluster. Firstly, the cluster emerges during December week 1, 2021 with 55 sites, 30 of which exhibit mutations characterizing B.1.1.529 (Omicron) but not in its BA.*-exclusive mutations, 13 sites exhibit BA.1-exclusive mutations and 13 exhibit Delta (B.1.617.2.*) reverse mutations (with Delta and Omicron sharing the nuc:28881 site), see Fig. 4 (C). Most of the sites exhibiting Omicron and BA.1 mutations (*>* 80%) have never established a relational pattern. Therefore, our critical cluster, when connected to Pango designations, corresponds to the emergence of Omicron and BA.1.

The critical cluster decreases in size to 12 during January week 3, 2022. Among the remaining 12 sites, 10 exhibit BA.1 mutations and 1 exhibits BA.2 mutation, see Fig. 4 (C). This indicates that Omicron outcompetes Delta.

The critical cluster increases in size to 36 during February week 1, 2022. The additional 24 sites exhibit BA.2 mutations. Besides the enlargement, we observe that the sites that previously exhibited BA.1 mutations experience reverse mutations. This indicates that BA.2 emerges and starts competing with BA.1, well before this manifests in terms of prevalence, see Fig. 4 (C).

The three alerts mark the emergence or disappearance of key variants (Delta, Omicron, BA.1 and BA.2). As in the case of the analysis of UK-data, the analysis of US-data provides a deeper insight into the constellation of characteristic mutations. It is worth pointing out that the site exhibiting mutation s:P681H in Alpha is linked to the site exhibiting s:N501Y [34, 35]. This linkage dissapears when this site is contained in Delta, where it exhibits the mutation s:P681R [36, 38, 39]. Furthermore, we can report that this linkage is recovered in Omicron, as both s:P681H and s:N501Y are present in Omicron.

#### 2.1.3 New York

We next present the more concurrent, prospective analysis based on New York GISAID data, starting from 02-04-2022 until 06-18-2022. In contrast to the previous two case studies, there exists no interpretation of the many Omicron sub-lineages reported. We analyze the New York data at a much shorter time scale, that is at a daily resolution, where each data point is averaged over three-consecutive-day periods. The analysis performed at this resolution allows us to put the applicability of the method to biased, as well as sparse data to the test.

Following the logic of the two previous case studies, we describe observations based on variant prevalence. We observe four key Omicron sub-lineages, BA.1, BA.2, BA.2.12.*, and BA.5. At a certain point in time, each respective lineage represents over 30% of the virus, see Fig. 5 (A). On 02-04-2022, BA.1 dominates (95%). A new sub-lineage BA.2.12.* emerges (5%) on 03-05-2022. On 04-23-2022, BA.1 represent around 5% and BA.2 and all of its sub-lineages, dominate (95%). BA.5 reaches 5% prevalence at 05-16-2022 and its prevalence increases, surpassing the BA.2 lineages. While BA.4 also surpasses a 5% prevalence in parallel with the emergence of BA.5. However, its prevalence increases slower than that of BA.5.

**Fig 5.**
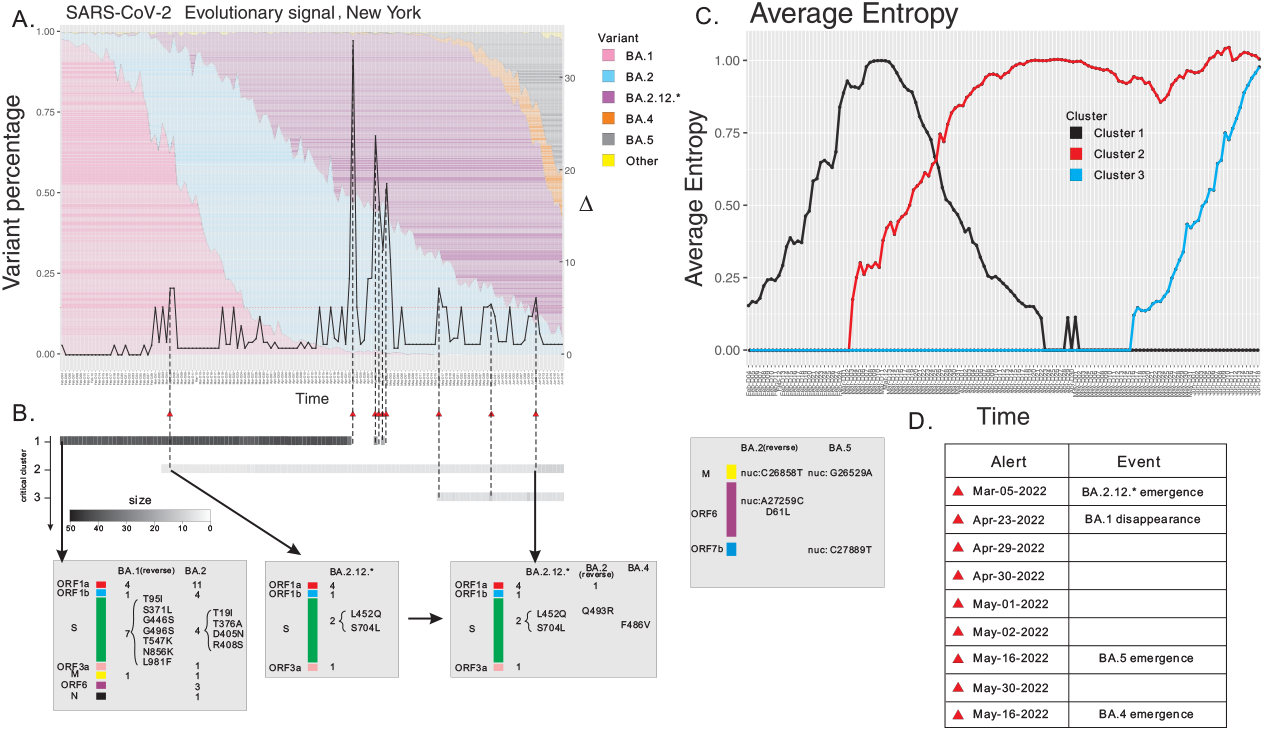
SARS-CoV-2 surveillance based at daily resolution of New York, USA data, 02-04-2022 through 06-18-2022. (A) The differential is displayed as a curve with its values on the y-axis. (B) The persistence of critical clusters, represented by bar codes, whose grey scale reflects the cluster size. Annotation of sites contained in critical clusters. We cross-reference the sites to characteristic mutations of reported variants. Reverse mutations are distinguished from mutations. Spike protein mutations as well as key mutations reported in the literature are listed. (C) Average entropy of sites contained in a critical cluster. (D) List of alerts (red triangles).

Our analysis issues nine alerts (A1)-(A9). (A1) is issued via a de novo cluster observed on 03-05-2022, coinciding with the emergence of BA.2.12.*. (A2)-(A6) are observed in the time frame between 04-23-2022 and 05-02-2022, coinciding with the disappearance of BA.1. It is worth pointing out that any data point in this analysis integrates sequence data of only three subsequent dates. This may induce a sampling bias not introduced at the larger time scales of the previous two case studies, where no such multiplicities are reported. (A7) heralds the emergence of BA.5 and (A8) is related to the enlargement of the cluster associated with BA.5. (A9) is related to the enlargement of the BA.2.12.*-cluster, see Fig. 5 (A).

The alerts (A1)-(A9) are associated with three critical clusters (C1)-(C3). (C1) is already present at the start of our time window, it consists of 38 sites at 02-04-2022, 13 of which exhibit BA.1 and 25 exhibit BA.2 mutations, respectively. None of these mutations are found in Omicron, its parent lineage B.1.1.529 [14]. The average entropy of (C1) exhibits a unimodal shape, due to the competition between BA.1. and BA.2. BA.1 and BA.2. exhibit shared as well as exclusive sites. Exclusive sites exhibiting BA.1 mutations are in fact reverse mutations. (C1) disappears completely on 05-02-2022.

The second cluster emerges on 03-05-2022, with 5 sites exhibiting mutations C11674T, T15009C, C21721T, T22917AG, C23673T. Specifically, C23673T and T22917A are non-synonymous nucleotide mutations that cause the amino acid mutations S704L and L452Q in the spike protein, respectively. The amino acid mutation S704L is reported as the bio-marker of BA.2.12^P^ and L452Q is reported as the bio-marker of BA.2.12.1^P^ [40–42]. Therefore, this critical cluster merges BA.2.12^P^ and BA.2.12.1^P^. Furthermore, the average entropy of the cluster is increasing, indicating that the prevalence of BA.2.12^P^ and BA.2.12.1^P^ is increasing. The cluster enlarges on 06-11-2022 by 4 additional sites, one of which exhibiting mutation Q493R in the spike protein found in BA.2, and one of which exhibiting the mutation F486V in the spike protein reported in BA.4 and BA.5 [40–42]. Thus Q493R and F486V are linked. The prevalence of Q493R is decreasing, indicating that it is reversed back to the wild type while F486V is increasing. This fact reflects the linkage is in fact due to the competition between BA.2 and BA.4&BA.5 3and the latter appear to outcompete BA.2.

The third critical cluster emerges on 05-16-2022 containing 7 sites, 5 of which exhibiting the mutations ORF6:D61L, nuc:C26858T, and nuc: A27259C in BA.2, and 2 exhibiting the mutations m:D3N and nuc:C27889T in BA.5 [40–42]. In particular, BA.4 and BA.5 share the same constellation of mutations on the spike protein, but m:D3N is a novel mutation of BA.5 that is not found in BA.4, Therefore, this cluster distinguishes BA.5 from BA.4. The cluster splits into two mutually exclusive blocks (m:D3N,nuc:C27889T) and (nuc:C26858T,nuc:A27259C,ORF6:D61L), with the prevalence of the former increasing and the prevalence of the latter decreasing.

In Table 1, we summarize our results on variants that become a threat i.e. a variant that assumes 50% of the viral population. We stipulate that an alert correctly identifies a variant if its critical clusters exhibit the characteristic mutations as designated by Pango [15].

**Table 1.** Motif-based alerts: we present the time an alert is issued, issue tickets #, see Pango designation website, bio-markers within the characteristic mutations based on Pango, gene-locations, Spike (Spike protein), ORF1a (Open Reading Frame 1a) and nuc (nucleotide mutation).

| Variants | Pango issue | Bio-markers | Regions | Alert-date |
| --- | --- | --- | --- | --- |
| B.1.1.7 (Alpha) | N/A | Spike: N501Y, P681H [34–36] | UK | Nov Week 1, 2020 |
| B.1.617.2 (Delta) | # 49 | Spike: E484Q, L452R [37, 43] | UK | Apr Week 3, 2021 |
| B.1.617.2.3/4 (AY.3/4) | # 121 | ORF1a: T3646A, I3731V [15, 44] | UK | Apr Week 3, 2021 |
| B.1.1.529 (Omicron) | # 343 | Spike: Q498R* [45, 46] | USA | Dec Week 1, 2021 |
| BA.2.12/BA.2.12.1 | # 499 | Spike: S704L, L452Q [40–42] | New York | 03-05-2022 |
| BA.4 | # 517 | Spike: F486V, nuc: G12160A [40–42] | New York | 06-11-2022 |
| BA.5 | # 517 | Spike: F486V, nuc: G26529A [40–42] | New York | 05-16-2022 |

We next derive an empirical notion of accuracy for our framework. To this end we define the number of positives (P) and negatives (N) as follows: each time interval contributes either one positive or negative, depending on whether or not a keystone event occurs. The number of alerts (PP) determines the number of non-alerts (PN).

We stipulate an alert is a true positive (TP) if it coincides with a positive, and a false positive (FP) otherwise. Similarly, a non-alert interval is considered a true negative (TN) if it coincides with a negative, and a false negative (FN) otherwise, i.e. where a keystone event occurs without triggering an alert. To quantify the accuracy of our prediction, we consider the resulting confusion matrix.

The alert-accuracy is then given by 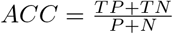 and in Table 2 we display the confusion matrices for three case studies, where the accuracy is the sum of the upper left and lower right entry divided by the sum of all entries of the respective matrix. In summary we obtain the alert-accuracies 88.6%, 100% and 99.3%, respectively.

**Table 2.** Alert accuracy of UK, USA and New York. We quantify the performance of our alerts on capturing keystone events, i.e. the emergence and disappearance of a dominating variant. Here *P* denotes the number of keystone events, *N* is the number of moments where no key events occur, *PP* (Predicted Positive) is the number of alerts and *PN* (Predicted Negative) is the number of intervals where no alert is triggered.

| | $PP$ | $PN$ |
| --- | --- | --- |
| $P$ | 3 | 0 |
| $N$ | 4 | 28 |
| (a) UK |  |  |
| $P$ | 3 | 0 |
| $N$ | 0 | 16 |
| (b) USA |  |  |
| $P$ | 8 | 0 |
| $N$ | 1 | 126 |
| (c) New York |  |  |

### 2.2 Geo-spatial robustness

In this subsection we provide a comparative study of alerts across different geographic locations.

We begin by curating SARS-CoV-2 genomic sequences collected during the Omicron emergence in 39 different geographic locations. The analysis covers 1 week before and 3 weeks after the emergence of Omicron. In order to have sufficient data, we considering all locations that collected at least 10^3^ sequences within the time window, at least 10^2^ of which were Omicron. By design, this curated data set represents the Delta-to-Omicron transition across the world.

For all 39 geographic locations, our framework issues an alert, coinciding with the advent of Omicron. We observe that the critical cluster of mutations inducing the alert is highly consistent across all different locations, see Fig. 6.

**Fig 6.**
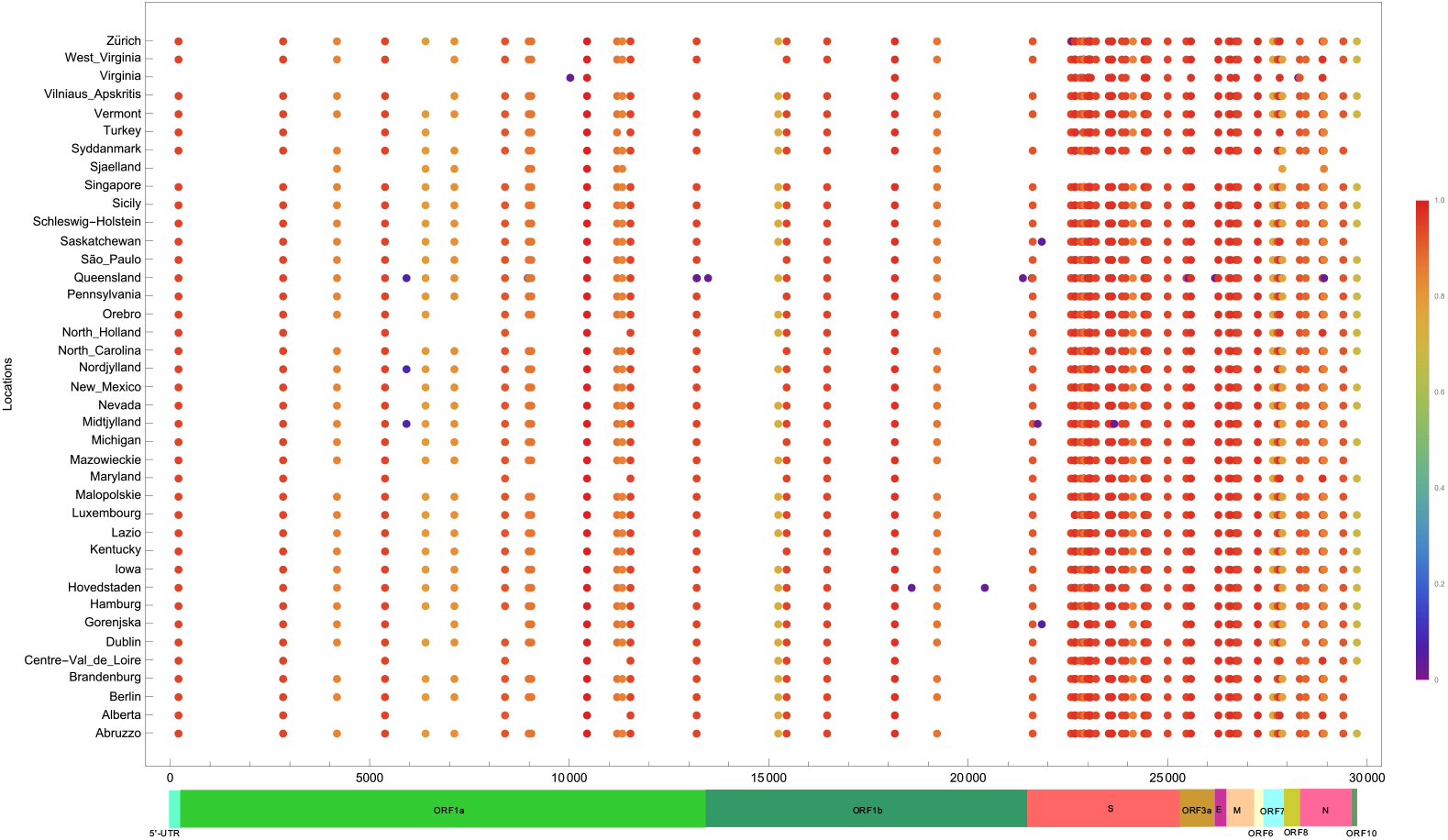
Critical clusters identified in SARS-CoV-2 evolution across different geographic locations. The x-axis denotes the genomic sites (SARS-CoV-2) and the y-axis denotes geographic location. Each dot represents a mutation of the critical cluster in the corresponding location. Columns are colored to represent the fraction of contributing locations. The bottom bar displays the genes annotation of the SARS-CoV-2 genome.

## 3 Discussion

We present a genomic surveillance system that alerts to potential threats in SARS-CoV-2. The system is based on a novel concept–the motif complex–that encapsulates the relational structure among sites, hidden within the MSA-data. By design, the motif complex is composed by *k*-ary relations among co-evolving sites and, compared to phylogeny-based methods, interprets the MSA in a substantially different way.

Current phylogenetic analysis, such as Pango and Nextstrain [13, 15], designate a new lineage per day on average. Each lineage, when designated, is considered to pose a potential threat. In difference to current approaches [9–15], where variants are considered to be a set of mutations, our surveillance framework, identifies “winning patterns” among sites within the MSA and as a result its alerts represent very different information. The motif complex views a variant as a collection of clusters of co-evolving sites. The rationale behind focusing on this is as follows: the probability of observing the emergence of co-evolutionary relations among multiple sites in a sequence of MSAs is extremely rare. Assuming that existing mutations have been positively selected, the formation of new linkages is indicative of better adaption–an event of relevance for the host.

A P-variant equals a collection of mutations and it is worth mentioning that this conceptualization has a specific drawback. There are two types of nucleotide modifications: mutations, which deviate from the wild type, and reverse mutations, which mutate back to the wild type. By construction P-variants consider only mutations. In contrast, the notion of a M-variant has intrinsically no reference to mutations or reverse mutations and is therefore impervious to this. Mutations and reverse mutations factor into the identification of the relational structure.

As for the relevance of reverse mutations: the critical cluster of co-evolving sites inducing an alert, typically splits into two mutually exclusive blocks, one block exhibiting mutations that coincide with those of the P-variant, and the other exhibiting reverse mutations. We emphasize that it is not just the *existence* of reverse mutations intrinsically, it is the *linkage* between reverse mutations and mutations. In fact, with one exception, BA.2.12.*, linked, reverse mutations are observed in the critical clusters that emerge during all alerts presented in this paper, see Fig. 1. Unlike the constellation of mutations used in order to characterize P-variants, the linkage among mutations and reverse mutations is, to the best of our knowledge, not investigated. The presence of linked reverse mutations in the critical clusters of all threats, however, signals its importance to facilitate a new co-evolutionary relation and is indicative of the occurrence of significant adaptation. We stipulate that the incorporation of these linkages is vital for biological analysis in general and vaccine development in particular.

We observe on May 16 an alert of an new cluster that exhibits a linkage between reverse mutations and mutations in the spike protein. Since this is a universal signature of threats, see Fig. 1, we predict –on May 16– BA.5 to be a threat, based on sequencing data from New York.

The notion of motif complex was introduced in [29] as a framework for the surveillance of SARS-CoV-2. In this paper we present a new alert criterion based on differential and cluster criticality. The differential quantifies the changes of the motif complex and allows to recognize when such significant changes occur. These mean that new co-evolutionary relations, representing better adaptation are realized. Cluster criticality involves persistence and dynamics. Critical clusters exhibit growth and shrinkage over time and dynamics is reflected by the Shannon entropy of their sites. In particular, the entropy is maximized when 50% of the sequences in the MSA exhibit the critical cluster.

We have shown in Section 2.1 that our alerts are able to capture keystone events in the evolutionary dynamics, including the emergence and disappearance of variants of concern. Compared to the concept of alert employed in [29], the alert introduced here is more sensitive, consistent and robust. This improvement facilitates a real-time, genomic surveillance.

In addition, our work hints towards a unified framework integrating bio-physics and some information theoretic augmentation of biology. The relational structure of an MSA is closely connected to that of molecular structures, such as RNA structures and proteins, where sites can be regarded as “nucleotides”, and the linkage among sites as a base pairing rule [47]. Incidentally, an RNA secondary structure is considered as a collection of non-crossing arcs in the upper half-plane [48], which is to say it is a collection of binary relations over sites. An RNA secondary structure thus constitutes a simple relational structure, in which all relations are equal and binary, namely

Watson-Crick and Wobble base pairs. An MSA comprised of sequences which all fold into a fixed RNA secondary structure will then induce a motif complex that allows one to retrieve these base pairs but also additional information that guarantees that the sequence folds correctly. This is studied in the context of consensus structure folding based on MSAs [49].

## 4 Materials and Methods

### 4.1 Analysis protocol

We outline the computational framework for our surveillance as follows:

1. Collecting the data. We obtain MSAs of SARS-CoV-2 genomes from GISAID and partition them into bins according to their sample collection time.
2. Constructing the motif complex. First we identify a subset of polymorphic sites having entropy *H*(*i*) *> h*_0_ in an MSA. Here *h*_0_ is a threshold for sites that exhibit sufficient nucleotide diversity. This process determines the 0-simplices of the motif complex. After selecting all 0-simplices, we approximate the 1-simplices by computing the *J* -distance between pairs of positions and approximate higher dimensional simplices via cluster analysis based on Highly Connected Subgraphs (HCS)-clustering [50]. For HCS clustering, the similarity graph is constructed using a threshold parameter *m*_0_, i.e., two sites are connected by an edge in the similarity graph if their co-evolution distance is *< m*_0_.
3. Computing the differential. Given two motif complexes, we construct a natural correspondence between their clusters. Our computation is based on a weighted bipartite graph having weights measured by the similarity (nearness) of two clusters, and is facilitated by the Hungarian algorithm of finding a perfect matching with a maximum total weight. We then integrate the displacement index between paired clusters to compute the differential, Δ, capturing the change of motif complex over time. This index will provide a signal for the surveillance of the virus.

We provide the technical details of each step below.

### 4.2 Motif complex

We recall the mathematical framework developed in [29] that utilizes the motif complex to detect co-evolutionary signals in a given MSA. The motif complex encapsulates the *k*-ary relationships within the columns of an MSA, which cannot be reduced to pairwise relations when these relationships are not transitive. In a natural way these *k*-ary relations for any *k* give rise to a weighted simplicial complex [51, 52], upon which the notion of co-evolutionary signals is derived.

An MSA is considered as a vector of random variables 𝕏 = (*X*_1_, *X*_2_, …, *X*_*n*_), where *X*_*i*_ is a random variable with four nucleotide states, *X*_*i*_ ∈ {*A, T, C, G*}. Each sequence in the MSA gives a sample 𝕏. A co-evolution pattern of the *i*th site is the distribution of *X*_*i*_ on the samples of MSA.

Suppose 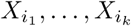 has a *k*-ary relationship (*M*_*k*_[*i*_1_, …, *i*_*k*_])_*k*_. Such a relationship can be considered to represent the existence of co-evolution between these *k* sites, which manifests via a variety of constellations of mutations. Instead of using a graph model whose edges represent pairwise relationships, the *k*-ary relationship (*M*_*k*_[*i*_1_, …, *i*_*k*_])_*k*_ gives rise to a simplicial complex of dimension (*k* − 1), which we refer to as the motif complex.

To investigate the motif complex, which is unknown to the observer, we utilize the MSA to obtain information about maximal motifs that govern the co-evolution. The “true” motif complex is approximated via the maximal collections of all sites having implicit connection, i.e., co-evolution. Specifically, co-evolutionary distances such as P-distance and J-distance [29] are utilized to quantify the coupling of pairs, and maximal motifs manifest as clusters.

### 4.3 Data preparation

SARS-CoV-2 whole genome data was collected from GISAID [10]. All collected sequences are aligned to the reference sequence collected from Wuhan, 2019 (GISAID ID: EPI ISL 402124). A multiple sequence alignment (MSA) was produced by MAFFT [21]. For the analysis of UK and USA data at a weekly resolution, sequences are partitioned into time bins according to their collected time, where week 1 equals day 1 to 7, week 2 equals day 8 to 15 and the following weeks are mapped accordingly. For the analysis of the New York data at a daily resolution, sequences are integrated over time bins containing three consecutive days.

### 4.4 Data reduction

SARS-CoV-2 genome contains roughly 3 × 10^4^ sites, whence considering all pairwise *J* -distances would be not only computationally intensive, but also provide irrelevant clusters. Namely, for the purpose of pattern inference, not all sites contain substantial information. This is evident when considering sites exhibiting no polymorphisms, whatsoever. Although one may hypothesize a relation between these sites, the MSA as a data structure does not provide any confirmation for such a hypothesis. Therefore, in a natural way, a threshold parameter, *h*_0_, comes into play, which controls the diversity of a site. To be specific, when the Shannon entropy of a site *i* has *H*(*i*) *> h*_0_, the site is taken into consideration for the construction of the motif complex, which is detailed below.

### 4.5 Motif complex approximation

Following [29] we approximate the motif complex as follows: we first utilize Shannon entropy to identify the active sites where selection induces evolutionary variation, and secondly, we employ *J* -distance to quantify those pairs of co-evolving sites. While sites with entropy greater than *h*_0_ can be viewed as 0-simplices, pairs of sites having *J* -distance greater than a threshold *m*_0_ form 1-simplices of the motif complex. This naturally induces a similarity graph for a fixed *m*_0_.

In order to reconstruct the maximal simplices of the motif complex, we perform the HCS-clustering on the similarity graph based on the *J* -distances between pairs of active sites. The key parameter for HCS-clustering is *m*_0_, as it determines when two sites are deemed to be co-evolving in the similarity graph. The clustering output provides us with the maximal sets of mutually co-evolving positions. The parameter *m*_0_ affects the motif complex and the effect of *m*_0_ on the differential is presented in the Supplementary Materials.

### 4.6 Cluster correspondence

We present a motif complex via a set of clusters *S* = {*C*_1_, …, *C*_*k*_}. The differential of two motif complexes, 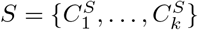 and 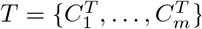, is derived from the similarity of two sets of clusters. To specify the latter, without loss of generality, we can assume *k* = *m*, otherwise, we enlarge *S* or *T*, by empty sets, respectively.

Suppose there is a bijective correspondence between 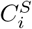 and 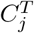 for 1 ≤ *i, j* ≤ *k*. We set 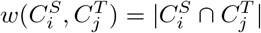. Now consider a weighted complete bipartite graph *G*, in which a vertex represents a cluster in either *S* or *T*. An edge is drawn between 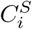 and 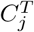 with weight 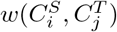. Then given a matching *M* of the bipartite graph *G*, the total weight of *M* is given by

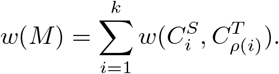

Here the matching *M* is represented as a permutation *ρ* over [*k*].

It is clear that two motif complexes observed at infinitesimally close points in time are essentially identical. In view of this, we stipulate that the clusters of two motif complexes at two consecutive points in time are to be matched by an *M* having maximum total weight in the weighted bipartite graph *G*. This problem can be solved by the Hungarian algorithm [53, 54] in polynomial time. The derived cluster correspondence provides in addition a trace of a cluster over time, facilitating an in-depth analysis of the time evolution of a cluster.

### 4.7 The differential of motif complexes

Given two motif complexes *S* and *T*, we have established the bijective correspondence between *S*-clusters and *T* -clusters as discussed in Sub-Section 4.6. For each pair of clusters in the correspondence, 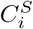 and 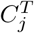, we define their *displacement index* as

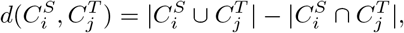

which quantifies difference between two configurations of clusters.

Given the cluster correspondence *M* between two motif complexes, we then define the differential in order to quantify the change of motif complex over time, by summing over the exponential of the displacement indices

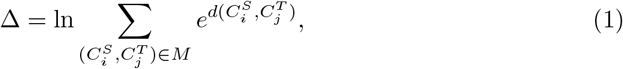

mimicking the construction of the partition function in statistical physics.

## Data Availability

All data produced in the present study are available upon reasonable request to the authors

## Acknowledgments

We want to thank Andrei Bura, Qijun He and Benjamin Reidys for their input on the manuscript. This work was supported by the VDH Grant PV-BII VDH COVID-19 Modeling Program VDH-21-501-0135.

