## Supplemental Materials for "Rapid threat detection in SARS-CoV-2"

### Supplementary Materials

**Simulation 1: a new variant from emergence to domination.** We first investigate a simulation starting with the population consisting of a single reference strain. The simulation is designed to highlight the behavior of the described system during the emergence of a new variant and the extinction of an old variant. It is simplified in such a way that factors such as mutational rate and population size can be ignored. The index strain has 20 nucleotides, and it dominates 100% of the population at the first generation and is used as a reference. Then in the second generation, a new variant, denoted by variant A, emerges and it takes over 10% of the population. Variant A has 10 mutations in sites 1 to 10 with respect to the reference strain. Then variant A takes over 10% more of the population in each of the following generations. At the 11-th generation, variant A completely dominates the population, and the reference strain becomes extinct.

In the first generation, each column in the MSA contains only one type of nucleotide, and thus the motif complex of the MSA consists of 20 isolated vertices. In the second generation when variant A emerges, all ten mutations present in the same sequence, sites 1 to 10 are correlated, exhibiting a co-evolution pattern. The last 10 sites remain the same as there are no mutations. Therefore, the motif complex in the second generation consists of a cluster of size 10, and 10 isolated vertices. The differential is given by $\Delta = 10$.

From generations 3 to 10, the increase of variant A in the population does not affect the co-evolving relation among sites 1 to 10, and the motif complex remains the same and $\Delta = 0$. In generation 11, the reference strain becomes extinct so that every column of the MSA contains only one type of nucleotide. The motif complex changes to 20 isolated vertices, having $\Delta = 10$ by definition. We show the $\Delta$ curve and the population constituent in Fig. 1 A. Here, we consider a signal when $\Delta > 5$.

We use a persistent bar code to illustrate the lifespan of a cluster with size $\geq 5$, see Fig. 1 B. We observe a cluster of size 10 from generation 2 to 10. The emergence and disappearance of this cluster correspond to the two signals triggered by $\Delta$ in generations 2 and 10 respectively. We point out that the magnitude of the signal $\Delta$ captures the degree of change of the motif complex.

We further study the relevancy of a cluster by considering the average entropy of sites contained in the cluster, denoted by $E(C_i)$. We show $E(C_1)$ in Fig. 1 C. Starting from generation 2, $E(C_1)$ increases when variant A emerges in generation 2 and takes over the population, and reaches its maximum $E(C_1) = 1$ when the population consists of 50% of variant A and 50% of the reference strain. After that the proportion of variant A keeps increasing, $E(C_1)$ decreases because the sites containing variant A mutations become less diverse. $E(C_1)$ reduces to 0 in the 11-th generation as the sites containing variant A mutations are no longer diverse. We conclude that cluster 1 is of interest from generation 2 to 10 as it persists in this time period. The average entropy of the cluster first increases, indicating that it becomes more and more relevant, and decreases after, indicating that its relevancy fades away.

In summary, our surveillance triggers signals via $\Delta > 5$. The signals indicate that there are significant changes in the motif complex. By tracking the lifespan of clusters that trigger the signal, we evaluate its relevance by the length of its lifespan, the size of the clusters, and the average entropy of the cluster. Long persistent clusters with large size and increasing/decreasing average entropy have increased potential to lend significant fitness and should be investigated further.

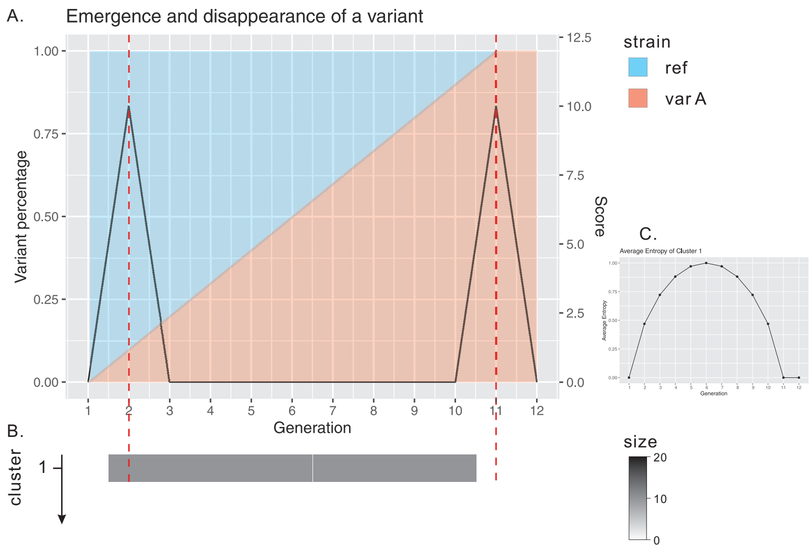


**Fig 1.** Evolutionary signal versus the proportion dynamics of strains in a population. LHS: An evolution simulation with two essential events: emergence of a new variant, and extinction of an old strain. The spike of the signal captures both of these events. RHS: An evolution simulation with two competing variants. The simulation has two extinction events. Our evolutionary signal is sensitive to both.

**Simulation 2: Competition of multiple variants.** The second simulation begins with a population composed of 20% reference strain, 40% variant A and 40% variant B, with all sequences having 20 nucleotides. Variant A has mutations from the reference in sites 1 to 10, while variant B has mutations from the reference in sites 11 to 20. Thus variant A and variant B differ in all 20 sites. Variant A and B increase by 10% in the second and third generation, respectively, resulting in the extinction of the reference strain by the third generation. After that, variant A takes over population by 10% more until variant B disappears in the 8-th generation.

When variants A, B and the reference strain are present in the population, the motif complex consists of two clusters: the first one contains sites 1 to 10, and the second cluster contains sites 11 to 20. The motif complex remains unchanged until the third generation when the reference strain disappears. As variant A and B mutations are mutually exclusive, sites 1 to 10 exhibit the same mutational pattern as sites 11 to 20, see Fig. 2. As a result, sites 1 to 20 become a cluster of size 20, which is merged from the previous two clusters of size 10. The cluster remains unchanged until generation 8 when variant B becomes extinct, giving $\Delta = 20$. We show the $\Delta$ curve and the population constituent in Fig. 2 A, the lifespan of the clusters in Fig. 2 B, and the average entropy of the clusters in Fig. 2 C. The average entropy of cluster 1 keeps decreasing, indicating that its relevancy is fading. The average entropy of cluster 1 suddenly drops in generation 3, as it is merged into cluster 1, and the relevancy of its mutations are accounted for in cluster 1.

The surveillance successively captures the extinction of the reference strain and variant B. We observe that the 8-th generation signal is twice as high as the one in the third generation. This is because the disappearance of a size 20 cluster changes the motif more significantly than the merge of two clusters of size 10. The proportional change of multiple variants in the population will not impact the motif complex.

**The effect of HCS-clustering parameter on the differential.**

Here we present a comparative analysis of $\Delta$ for different choices of the HCS-clustering parameter $m_0$, based on the USA SARS-CoV-2 genomic data starting from 11-08-2021 to 03-07-2022 on a weekly basis. By design, the parameter $m_0$ controls the graph connectivity in the HCS clustering.

Fig. 3 shows that the differential $\Delta$ is robust, across different choices of the clustering threshold $m_0$. Since the differential measures the change of motif complex, the result first indicates that our motif complex approximation for an MSA, is stable with respect to different choices of $m_0$. The co-evolutionary signals detected by our method truly reflect the linkage of sites in the MSA. It also suggests that the evolution dynamics of variants captured by $\Delta$ is not affected by the change of $m_0$.

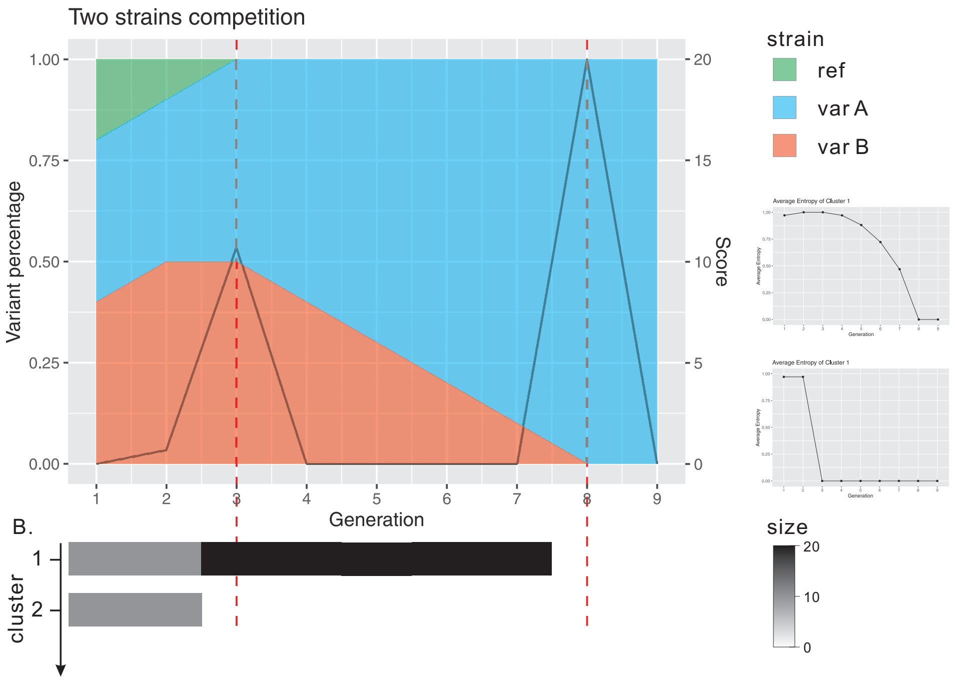


**Fig 2.** Evolutionary signal versus the proportion dynamics of strains in a population. LHS: An evolution simulation with two essential events: emergence of a new variant, and extinction of an old strain. The spike of the signal captures both of these events. RHS: An evolution simulation with two competing variants. The simulation has two extinction events. Our evolutionary signal successively captures both.

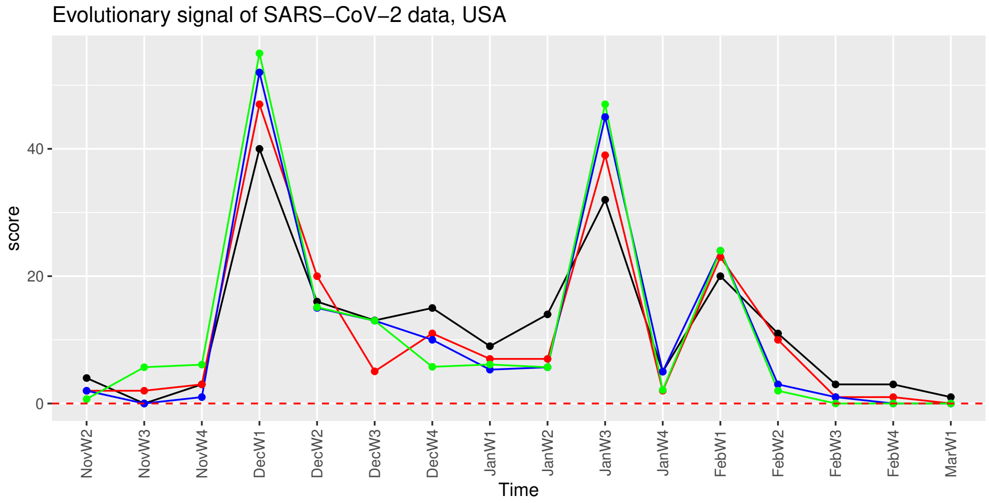


**Fig 3.** The differential $\Delta$ for varying $m_0$, for SARS-CoV-2 genomic data (USA) between November week 2, 2021 and March week 1, 2022 (weekly). The parameter $h_0 = 0.01$ is fixed and $m_0$ is varied: $m_0 = 0.3$ (black), 0.4 (red), 0.5 (blue), 0.6 (green). The $y$-axis represents the differential, and $x$-axis displays the time.
